# Clinical severity of COVID-19 patients admitted to hospitals during the Omicron wave in South Africa

**DOI:** 10.1101/2022.02.22.21268475

**Authors:** Waasila Jassat, Salim S Abdool Karim, Caroline Mudara, Richard Welch, Lovelyn Ozougwu, Michelle J. Groome, Nevashan Govender, Anne von Gottberg, Nicole Wolter, Milani Wolmarans, Petro Rousseau, DATCOV author group, Lucille Blumberg, Cheryl Cohen

## Abstract

**Background:** Clinical severity of patients hospitalised with SARS-CoV-2 infection during the Omicron (fourth) wave was assessed and compared to trends in the D614G (first), Beta (second), and Delta (third) waves in South Africa.

**Methods:** Weekly incidence of 30 laboratory-confirmed SARS-CoV-2 cases/100,000 population defined the start and end of each wave. Hospital admission data were collected through an active national COVID-19-specific surveillance programme. Disease severity was compared across waves by post-imputation random effect multivariable logistic regression models. Severe disease was defined as one or more of acute respiratory distress, supplemental oxygen, mechanical ventilation, intensive-care admission or death.

**Results:** 335,219 laboratory-confirmed SARS-CoV-2 admissions were analysed, constituting 10.4% of 3,216,179 cases recorded during the 4 waves. In the Omicron wave, 8.3% of cases were admitted to hospital (52,038/629,617) compared to 12.9% (71,411/553,530) in the D614G, 12.6% (91,843/726,772) in the Beta and 10.0% (131,083/1,306,260) in the Delta waves (p<0.001). During the Omicron wave, 33.6% of admissions experienced severe disease compared to 52.3%, 63.4% and 63.0% in the D614G, Beta and Delta waves (p<0.001). The in-hospital case fatality ratio during the Omicron wave was 10.7%, compared to 21.5%, 28.8% and 26.4% in the D614G, Beta and Delta waves (p<0.001). Compared to the Omicron wave, patients had more severe clinical presentations in the D614G (adjusted odds ratio [aOR] 2.07; 95% confidence interval [CI] 2.01-2.13), Beta (aOR 3.59; CI: 3.49-3.70) and Delta (aOR 3.47: CI: 3.38-3.57) waves.

**Conclusion:** The trend of increasing cases and admissions across South Africa’s first three waves shifted in Omicron fourth wave, with a higher and quicker peak but fewer admitted patients, who experienced less clinically severe illness and had a lower case-fatality ratio. Omicron marked a change in the SARS-CoV-2 epidemic curve, clinical profile and deaths in South Africa. Extrapolations to other populations should factor in differing vaccination and prior infection levels.

## INTRODUCTION

The fifth SARS-CoV-2 variant of concern, Omicron (B.1.1.529 lineage), was first publicly announced in South Africa on 25 November 2021 (1). Within days, Omicron led to a resurgence of SARS-CoV-2 cases in South Africa and several other countries (2).

Genomic sequencing revealed that 86% (n=1,353) and 99% (n=1,361) of sequenced SARS-CoV-2 samples nationally in South Africa were Omicron in November and December respectively (3). The proportion of ThermoFisher TaqPath COVID-19 real-time reverse transcription polymerase chain reaction (rRT-PCR) positive tests with S-gene target failure, a marker of Omicron (4), was 2% (n=11), 95% (n=10,536) and 98% (n=19,174) in October, November and December respectively (5). Phylogenetic surveillance showed that the predominant variants (>90% of viral sequences) during South Africa’s first, second and third waves were the ancestral strain with a D614G mutation, Beta and Delta respectively (3).

The mutations identified in Omicron suggested that it would likely be highly transmissible with immune escape but provided no indication on whether it would have a different clinical severity profile compared to previous variants (1). Clinical severity of COVID-19 is influenced by several factors besides virulence of the viral variant, including age, sex, race, co-morbidities, vaccination status, immunity from previous SARS-CoV-2 infection and early therapy/prophylaxis.

The South African COVID-19 vaccination programme began with healthcare workers from February 2021. It subsequently expanded to adults older than 60 years in May 2021 and then progressively over time to other age groups until adolescents 12-17 years were included in mid-October 2021. As a result, vaccination coverage was low during the Delta wave but by 17 November 2021 before the Omicron wave started, 35% of South Africa’s adult population were fully vaccinated with either two doses of BNT162b2 or one dose of Ad26.CoV2.S vaccine (8). A third dose of BNT162b2 and a second dose of Ad26.CoV2.S only became available at the tail end of the Omicron wave. Further, a substantial number of people have been infected with SARS-CoV-2 during each wave. SARS-CoV-2 seroprevalence was 47% in blood donors in South Africa after the second wave (6), and 73% in a community-based survey in Gauteng province after the third wave (7). Early treatment with Remdesivir or monoclonal antibodies is not widely available and is infrequently used in South Africa.

We assessed the clinical severity of patients hospitalised with laboratory-confirmed SARS-CoV-2 infection during the Omicron wave and whether it differed from the D614G, Beta and Delta waves in South Africa.

## METHODS

Data on real-time reverse transcription polymerase chain reaction (rRT-PCR) and antigen positive SARS-CoV-2 cases were collated daily from laboratory reports (9) while data on COVID-19 hospital admissions were collected through DATCOV, an active surveillance programme established specifically for COVID-19 (10). Secondary data analysis was conducted using the DATCOV national hospital surveillance database between 5 March 2020 and 22 January 2022. DATCOV surveillance collects data on all individuals with a positive SARS-CoV-2 rRT-PCR test or antigen test, with a confirmed duration of stay in hospital of one full day or longer, regardless of reason for admission. This included patients who had COVID-19 symptoms, were admitted for isolation, acquired nosocomial COVID-19 infection, or tested positive incidentally when admitted for other reasons. Incidental positive SARS-CoV-2 tests have been noted to occur mainly during the initial period when cases are rising in all four waves in South Africa. Some of the patients admitted in a wave may have been admitted in the previous waves; these repeat admissions (more than 90 days after the first positive SARS-CoV-2 test) were included in the analysis. Incidence risks were calculated using Statistics South Africa mid-year population figures for 2020 (11).

The wave periods were defined from the week the country crossed a weekly incidence risk of 30 cases per 100,000 persons at the start and end of the waves (12)(13). The Omicron-dominated wave crossed the weekly incidence risk threshold in the last week of November 2021. The start of each of these four wave periods selected also correlated with the majority of cases being due to the D614G, Beta, Delta and Omicron variants respectively (3). The full wave periods were included for all four waves.

- Wave 1: week 24 (2020)- week 34 (2020) (7 June-22 August 2020; 76 days)
- Wave 2: week 47 (2020)- week 5 (2021) (15 November 2020-6 February 2021; 83 days)
- Wave 3: week 19 (2021)- week 37 (2021) (9 May-18 September 2021; 132 days)
- Wave 4: week 47 (2021)- week 3 (2022) (21 November 2020-22 January 2022; 62 days)

Analysis of severity was restricted to admissions that had already accumulated outcomes and all patients still in-hospital or transferred to other hospitals without final outcomes were excluded, because they remained at risk of still developing severe outcomes including death. Descriptive statistics were used to describe the trends in cases, admissions, severe disease and death over the equivalent periods of the D614G (first), Beta (second), Delta (third) and Omicron (fourth) waves.

Post-imputation random effect (on admission facility) multivariable logistic regression models were used to compare severe disease between the waves. To account for incomplete or missing data on selected variables, we used multivariate imputation by chained equation (MICE) and generated ten complete imputed datasets that were used for subsequent analyses. Variables analysed using MICE included race and comorbidities, where up to a third of the data were missing. Complete or near-complete variables included in the imputation process were age, sex, province, health sector (i.e. public or private), severity and in-hospital outcome (i.e. discharged alive or died).

Severe disease was defined as one or more of the following: development of acute respiratory distress syndrome, receipt of oxygen or invasive mechanical ventilation, treatment in high-care or intensive-care units (ICUs) or death, using a modified definition based on the recommendations from the World Health Organization (WHO) Clinical Platform External Clinical Advisory Group (14). Age, sex, race, presence of a comorbidity (which included hypertension, diabetes, chronic cardiac disease, chronic kidney disease, asthma/chronic pulmonary disease, malignancy, HIV or tuberculosis), type of health sector (private or public) and province were included in the model to assess the relationship between each wave period and severity in SARS-CoV-2 positive patients admitted to hospital. The presence of obesity, while important, was excluded from the analysis due to poor completeness of this variable. The statistical analysis was implemented using Stata 15 (Stata Corp®, College Station, Texas, USA). We followed STROBE guideline recommendations.

## RESULTS

South Africa experienced four distinct waves of SARS-CoV-2 infections, with approximately three-month periods of low transmission between each wave (Figure 1). The 7-day moving average of daily cases had a peak that was higher in each successive peak of the four waves. While the wave duration increased across the first three waves, it declined in the fourth wave. The number of SARS-CoV-2 positive cases identified during each wave was 553,530, 726,772, 1,306,260 and 629,617 in the D614G, Beta, Delta and Omicron waves respectively (Table 1). Unlike the pattern observed in the prior waves, the rise in cases during the Omicron wave was not accompanied by a concomitant rise in hospital admissions (Figure 1); instead the peak in admissions was lower and of shorter duration. The percent of cases admitted was 12.9% (71,411/553,530) during the D614G wave, 12.6% (91,843/726,772) during the Beta wave and 10.0% (131,083/1,306,260) during the Delta wave compared to 8.3% (52,038 /629,617) during the Omicron wave (p<0.001).

**Table 1:** Summary of SARS-CoV-2 cases, COVID-19 admissions and in-hospital deaths in the D614G (7 June-22 Aug 2020), Beta (15 Nov 2020-6 Feb 2021), Delta (9 May-18 Sep 2021) and Omicron waves (21 Nov 2021-22 Jan 2022), South Africa

| Variant wave | Number of SARS-CoV-2 positive cases | Incidence of SARS-CoV-2 positive cases per 100,000 persons | Percent (Number) of cases admitted to hospital | Percent (Number) of admitted cases with an outcome who died in hospital (n) |
| --- | --- | --- | --- | --- |
| All ages (population 59,622,350; fully vaccinated 25.0%<sup>†</sup>) |  |  |  |  |
| D614G | 553530 | 928.4 | 12.9 (71411) * | 21.5 (15144) * |
| Beta | 726772 | 1219.0 | 12.6 (91843) * | 28.8 (26032) * |
| Delta | 1306260 | 2190.9 | 10.0 (131083) * | 26.4 (33947) * |
| Omicron | 629617 | 1056.0 | 8.3 (52038) | 10.7 (4907) |
| Age ≤5 years (population 5,743,450, fully vaccinated 0%) |  |  |  |  |
| D614G | 6280 | 109.3 | 12.9 (812) * | 5.1 (41) * |
| Beta | 8522 | 148.4 | 15.1 (1284) * | 4.5 (56) * |
| Delta | 19019 | 331.1 | 14.7 (2797) * | 4.0 (109) * |
| Omicron | 14050 | 244.6 | 25.4 (3568) | 2.1 (67) |
| Age 6-19 years (population 16,082,084, fully vaccinated 1.5%<sup>†</sup>) |  |  |  |  |
| D614G | 41829 | 260.1 | 3.7 (1550) * | 3.9 (59) * |
| Beta | 56873 | 353.6 | 2.7 (1517) * | 4.8 (70) * |
| Delta | 188689 | 1173.3 | 2.3 (4359) * | 2.9 (121) * |
| Omicron | 68778 | 427.7 | 5.7 (3899) | 1.8 (62) |
| Age 20-39 years (population 20,684,164; fully vaccinated 22.9%<sup>†</sup>) |  |  |  |  |
| D614G | 223169 | 1078.9 | 6.7 (15039) * | 7.1 (1051) * |
| Beta | 271440 | 1312.3 | 5.8 (15658) * | 10.3 (1581) * |
| Delta | 483252 | 2336.3 | 5.0 (24241) * | 9.3 (2193) * |
| Omicron | 273902 | 1324.2 | 6.2 (17027) | 4.3 (649) |
| Age 40-59 years (population 11,686,162; fully vaccinated 46.0%<sup>†</sup>) |  |  |  |  |
| D614G | 208211 | 1781.7 | 14.2 (29504) * | 16.9 (4933) * |
| Beta | 258981 | 2216.1 | 13.9 (35958) * | 23.1 (8182) * |
| Delta | 437093 | 3740.3 | 11.2 (48909) * | 23.1 (11097) * |
| Omicron | 188486 | 1612.9 | 6.5 (12259) | 11.2 (1199) |
| Age ≥60 years (population 5,426,490; fully vaccinated 57.8%<sup>†</sup>) |  |  |  |  |
| D614G | 74041 | 1364.4 | 33.1 (24506) * | 37.5 (9266) * |
| Beta | 130956 | 2413.3 | 28.6 (37426) * | 43.9 (16469) * |
| Delta | 178207 | 3284.0 | 28.5 (50777) * | 40.9 (20872) * |
| Omicron | 84401 | 1555.4 | 18.1 (15285) | 22.2 (2008) |

<sup>†</sup> Vaccination coverage data for Omicron wave is from 14 November 2021 (National Department of Health. Latest Vaccine Statistics. https://sacoronavirus.co.za/latest-vaccine-statistics/)
\* p<0.001 comparing Omicron wave to the D614G, Beta and Delta waves

**Figure 1:**
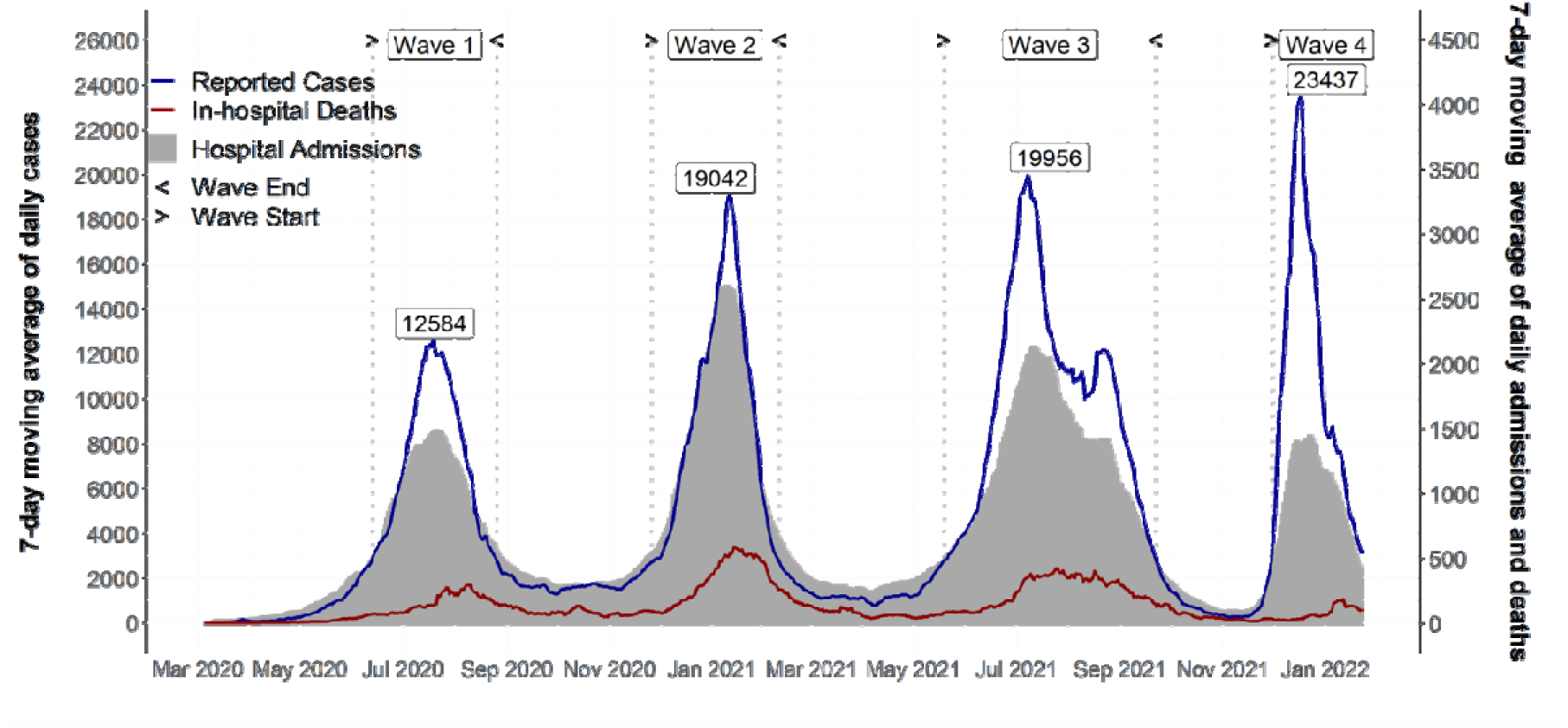
7 day moving average of SARS-CoV-2 cases, COVID-19 admissions and in-hospital deaths in South Africa, 5 March 2020-22 January 2022.

For the Omicron wave, clinical outcomes were known for 88.3% of the 52,038 SARS-CoV-2 positive patients admitted to hospital as of 22 January 2022; the remainder were still in hospital and did not yet have a documented in-hospital outcome. (Outcomes were unknown for 1.4% (987/71,411), 1.7% (1,533/91,843) and 1.9% (2,525/131,803) of patients admitted in the D614G, Beta and Delta waves.) In patients with known clinical outcomes, 52.3% (36,837/70,424) in the D614G wave, 63.4% (57,247/90,310) in the Beta wave and 63.0% (81,040/128,558) in the Delta wave compared to 33.6% (15,421/45,927) in the Omicron wave met the criteria for severe disease (p<0.001) (Table 2). The proportion of patients requiring supplemental oxygen was lower during the Omicron wave (10,565/45,927; 23.0%) compared to the D614G (25,890/70,424; 36.8%), Beta (43,235/90,310; 47.9%) and Delta (61,318/128,558; 47.7%) waves (p<0.001) (Figure 2). Median hospital stay was lower in the Omicron wave, 4 days [IQR 2-8 days], compared to 6 days [IQR: 3-11], 6 days [IQR: 3-10], 6 days [IQR: 3-11] in the D614G, Beta and Delta waves respectively (p<0.001) (Table 2). The in-hospital case fatality ratio during the Omicron wave (Table 1) was 10.7%, compared to 21.5%, 28.8% and 26.4% in the D614G, Beta and Delta waves respectively (p<0.001).

**Table 2:** Indicators of disease severity among SARS-CoV-2 positive cases admitted in the D614G (7 June-22 Aug 2020), Beta (15 Nov 2020-6 Feb 2021), Delta (9 May-18 Sep 2021) and Omicron waves (21 Nov 2021-22 Jan 2022), South Africa

| Variant wave | Number of cases admitted to hospital with known outcome | Median length of stay in days (IQR) | Percent (Number) of admitted cases who received supplemental oxygen (n) | Percent (Number) of admitted cases who were treated in ICU (n) | Percent (Number) of admitted cases who had severe disease (n) |
| --- | --- | --- | --- | --- | --- |
| All ages |  |  |  |  |  |
| D614G | 70424 | 6 (3-11) * | 36.8 (25890) * | 15.8 (11125) * | 52.3 (36837) * |
| Beta | 90310 | 6 (3-10) * | 47.9 (43235) * | 12.8 (11597) * | 63.4 (57247) * |
| Delta | 128558 | 6 (3-11) * | 47.7 (61318) * | 14.6 (18812) * | 63.0 (81040) * |
| Omicron | 45927 | 4 (2-8) | 23.0 (10565) | 6.3 (2872) | 33.6 (15421) |
| Age <5 years |  |  |  |  |  |
| D614G | 798 | 4 (2-8) * | 14.0 (112) ** | 9.1 (73) * | 24.8 (198) * |
| Beta | 1249 | 4 (2-8) * | 21.1 (263) * | 7.8 (97) * | 29.0 (362) * |
| Delta | 2732 | 3 (2-6) * | 18.6 (509) * | 6.7 (184) * | 28.1 (767) * |
| Omicron | 3242 | 3 (2-5) | 13.3 (432) | 3.9 (127) | 19.3 (627) |
| Age 5-19 years |  |  |  |  |  |
| D614G | 1529 | 5 (2-9) * | 13.1 (200) * | 7.1 (108) * | 20.8 (318) * |
| Beta | 1471 | 4 (2-9) * | 20.9 (308) * | 5.4 (80) * | 29.0 (427) * |
| Delta | 4212 | 5 (2-9) * | 18.2 (768) * | 4.9 (206) * | 25.1 (1056) * |
| Omicron | 3503 | 3 (2-5) | 11.0 (384) | 3.4 (118) | 16.9 (593) |
| Age 20-39 years |  |  |  |  |  |
| D614G | 14809 | 5 (2-10) * | 22.5 (3330) * | 8.7 (1293) * | 31.5 (4672) * |
| Beta | 15361 | 5 (3-9) * | 34.4 (5290) * | 7.7 (1177) * | 42.6 (6547) * |
| Delta | 23700 | 5 (3-9) * | 32.2 (7641) * | 8.7 (2059) * | 41.0 (9718) * |
| Omicron | 15259 | 3 (2-7) | 13.9 (2121) | 3.3 (502) | 21.0 (3202) |
| Age 40-59 years |  |  |  |  |  |
| D614G | 29120 | 7 (4-11) * | 39.3 (11441) * | 17.0 (4937) * | 53.3 (15515) * |
| Beta | 35426 | 6 (4-10) * | 49.6 (17574) * | 14.9 (5290) * | 63.5 (22508) * |
| Delta | 48010 | 7 (4-11) * | 51.7 (24817) * | 17.8 (8553) * | 66.3 (31854) * |
| Omicron | 10703 | 5 (2-8) | 25.5 (2728) | 6.8 (723) | 37.1 (3975) |
| Age ≥60 years |  |  |  |  |  |
| D614G | 24168 | 7 (3-13) * | 44.7 (10807) * | 19.5 (4714) * | 66.8 (16134) * |
| Beta | 36803 | 6 (3-11) * | 53.8 (19800) * | 13.5 (4953) * | 74.5 (27403) * |
| Delta | 49904 | 7 (4-12) * | 55.3 (27583) * | 15.7 (7810) * | 75.4 (37645) * |
| Omicron | 13220 | 5 (3-9) | 37.1 (4900) | 10.6 (1402) | 53.1 (7024) |

\* p<0.001; \*\* p>0.05 comparing Omicron wave to the D614G, Beta and Delta waves

**Figure 2:**
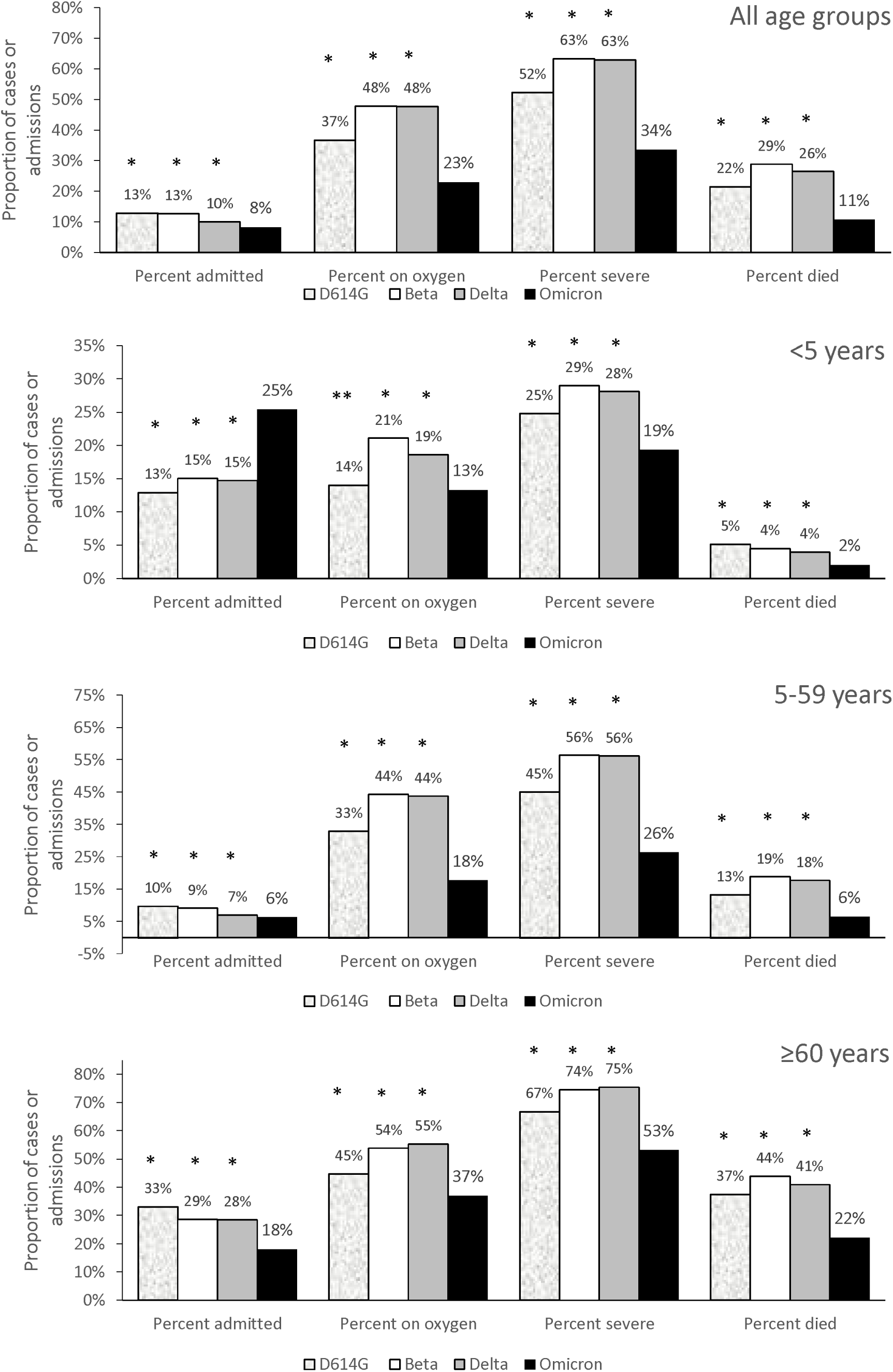
Percent of cases admitted, percent of admissions who received supplementary oxygen, with severe disease, and in-hospital deaths, for individuals of all ages (**Figure 2a**), <5 years (F**igure 2b**), 6-59 years (**Figure 2c**) and ≥60 years (**Figure 2d**) in the D614G (7 June-22 Aug 2020), Beta (15 Nov 2020-6 Feb 2021), Delta (9 May-18 Sep 2021) and Omicron waves (21 Nov 2021-22 Jan 2022), South Africa. * p<0.001; ** p>0.05 comparing Omicron wave to the D614G, Beta and Delta waves

Children and adolescents aged <20 years constituted 14.3% (7,467/52,038) of total admissions in the fourth wave, compared to 3.3% (2,362/71,411), 3.0% (2,801/91,843), and 5.5% (7,156/131,083) in the D614G, Beta and Delta waves. In children <5 years, 25.4% (3,568/14,050) of laboratory-confirmed cases were admitted in the Omicron wave compared to 14.7% (2,797/19,019) in the Delta wave, but the number of children <5 years hospitalised was similar in both waves and were in turn higher than the number of children hospitalised in the D614G and Beta waves (Table 1). The proportion of hospitalised individuals aged <5 years who had severe disease was lower in the Omicron wave (627/3,242; 19.3%) compared to the D614G (198/798; 24.8%), Beta (362/1,249; 29.0%), and Delta (767/2,732; 28.1%) waves (p<0.001).

On multivariable analysis, compared to patients admitted in the Omicron wave, patients were more likely to have severe disease if admitted in the D614G wave (adjusted odds ratio [aOR] 2.07; 95% confidence interval [CI] 2.01-2.13), Beta wave (aOR 3.59; CI 3.49-3.70) and Delta wave (aOR 3.47; CI 3.38-3.57) (Table 3). Other factors associated with severe disease in this patient population were older age, male sex, Indian compared to white race, presence of a comorbidity and the province of admission.

**Table 3.**
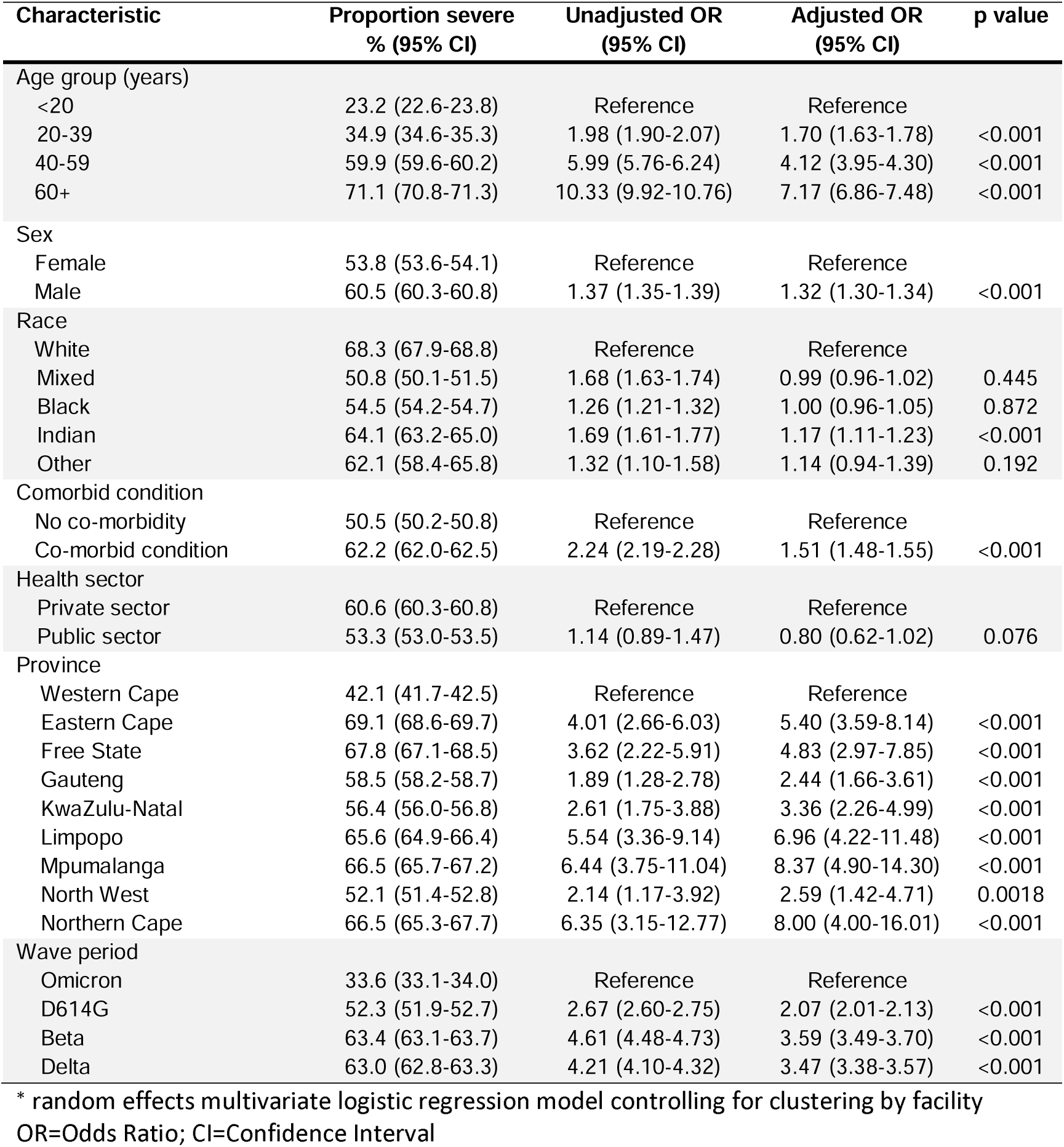
Factors associated with severe disease among SARS-CoV-2 positive hospitalised patients in the D614G (7 June-22 Aug 2020), Beta (15 Nov 2020-6 Feb 2021), Delta (9 May-18 Sep 2021) and Omicron waves (21 Nov 2021-22 Jan 2022), South Africa. (univariate and multivariable analysis implemented on the imputed dataset) (N=335,219)*

## DISCUSSION

The Omicron wave had a lower proportion of SARS-CoV-2 cases admitted to hospital while those admitted had shorter hospital stays and less severe illness, with fewer requiring oxygen or intensive care treatment compared to the D614G, Beta or Delta waves in South Africa. Both disease severity and in-hospital case fatality ratio were at least 2-fold higher in the three previous waves compared to the Omicron wave. The change in disease severity was more marked in adults than children.

The number of adults aged ≥20 years admitted to hospital was substantially lower in the Omicron wave compared to past waves leading to lower clinical burdens in health care services. The patients admitted during the Omicron wave placed less demand on oxygen supplies, ventilators and ICU beds than patients in the previous waves. Early reports from other countries also suggest reduced severity among hospital admissions in the Omicron wave (15,16).

The admission rate was higher in the largely unvaccinated <20 years age group, especially in children aged <5 years, in the Omicron wave compared to the first three waves. However, the admitted children <5 years had less severe illness in the Omicron wave compared to the first three waves.

Early reports from the United Kingdom also indicate an increased admission rate but decreased severity among children in the Omicron wave (17). Possible reasons for the higher admission rate in children could be that higher transmissibility led to more infections in children, more incidental infection amongst children admitted for other reasons, or their lower rates of prior infection (7) and/or vaccination (8).

The lower admission rates and less severe infections in admitted patients during the Omicron wave are most likely to be due to a combination of a less virulent virus, immunity from vaccination and prior infection(s), especially the large numbers of vaccinated individuals who had prior infection and so have “hybrid immunity” (18). Tissue-based studies showed that Omicron infects the cells of the bronchus more efficiently but alveolar cells of the lungs less efficiently than the Delta variant (19,20). The lower virulence of Omicron has also been demonstrated in animal models; mice have less severe disease with Omicron (21-23). The lower virulence of Omicron, which at least partially accounts for the less severe infections observed in the Omicron waves in several countries, could be contributing to its higher infection rates as infectious individuals remain clinically well and mobile thereby continuing to spread the virus within the community. The higher efficiency of upper airway infection may lead to children becoming symptomatic more often due to their smaller airways becoming more readily congested.

Reinfections with Omicron in those with prior infection are high (25). While prior infection may not prevent symptomatic breakthrough infection, it may generate T-cell responses that provide protection from severe disease (26,27), thereby contributing, at least partially to the observed high infection rate but lower severity with Omicron. South Africa experienced a particularly severe wave of Delta infection leading to a large increase in seroprevalence following the Delta-driven third wave. If prior infection with the Delta variant specifically provides some T-cell immunity that protects against severe disease from Omicron infection, this could be a contributor to, the less severe infections observed in the Omicron-driven fourth wave.

While SARS-CoV-2 vaccine effectiveness in preventing symptomatic infection has been impacted by variants (28), vaccines remain effective in reducing the risk of severe disease (29), including against Omicron (30). Since vaccination coverage was higher before the Omicron wave in individuals aged above 60 years (58%), it may have made an important contribution to the lower severity of Omicron infections, especially in the elderly. But vaccination cannot fully account for the markedly lower numbers of severe infections in 20-39 year-old individuals, as less than a quarter of this age group was vaccinated. Further, since vaccinations started only in mid-2021 in South Africa, a substantial number of vaccinated individuals have hybrid immunity which retains higher Omicron neutralisation than vaccination alone (31).

One of the limitations of this study is that clinical outcomes are not known for 11% of patients in the Omicron wave as it ended at the time of analysis and some patients are still in hospital. As clinical outcomes have varied little over the course of the Omicron wave, it is not anticipated that these results will change substantially when the outstanding clinical outcomes are added. Additionally, testing strategies for determining cases have changed over time, though most testing during the waves has focused on testing those with symptoms and those with exposure. While the criteria for hospital admission with Covid-19 may have changed over time, they have been minimal over the last six months when both the Delta and Omicron waves occurred.

This study has some data limitations as well. Firstly, disease severity relies on clinical parameters like oxygen and ventilation treatment and not laboratory parameters, although oxygen is usually initiated based on an objectively measured oxygen saturation. Secondly, the incompleteness of reporting in DATCOV and missing values in some patient data may under-estimate severity, but the completeness of reporting is unlikely to have changed substantially over the four waves. Thirdly, while the dataset did not have individual-level data on infecting lineage for cases included in this analysis, each of the four waves included in this study had a predominant variant that allows for wave period to be used as a proxy for dominant variant. During the fourth wave, genomic sequencing as well as S-gene target failure showed that over 95% of circulating viruses were Omicron. Fourthly, DATCOV contains incomplete data on prior SARS-CoV-2 infection and vaccination status, which limits exploration at individual patient level of their potential roles in lower disease severity observed. Infection and re-infection are substantially under-ascertained due to the high proportion of asymptomatic infections, especially in the large number of young people in South Africa. Data on COVID-19 hospital admissions are collected at health service level by clinicians and nurses and contains limited data on self-reported vaccination status; vaccination data is collected in a different system and linkage of the two data systems is still underway. This limitation has highlighted the need for the creators of the different surveillance datasets to ensure compatibility and potential for integration.

## CONCLUSION

The trend of increasing cases and admissions across South Africa’s first three waves shifted in the Omicron fourth wave. The Omicron wave was characterised by a higher and quicker peak but fewer admitted patients, who experienced less clinically severe illness and had a lower case-fatality ratio. Omicron marked a change in the SARS-CoV-2 epidemic curve, clinical profile and deaths in South Africa. Early reports from several other countries also indicate less severe disease with Omicron, despite the differences in population structure, co-morbidity prevalence, prevalence of prior infection and vaccination coverage. Since each of the five variants of concern have evolved independently of each other, it remains speculative in the absence of data as to whether the next variant will follow the trend of greater severity seen progressively with the first three waves or follow the low severity observed with Omicron. Data from a well-developed surveillance system for variant identification and hospital admissions is essential as part of pandemic preparedness to rapidly investigate the impact of new SARS-CoV-2 variants and viruses causing future pandemics.

## Data Availability

The dataset analysed for the manuscript is available upon reasonable request. The data dictionary is available at request to the corresponding author

## ACKNOWLEDGEMENTS

We acknowledge the National Institute for Communicable Diseases (NICD) team responsible for reporting test, case and hospitalisation data. Thanks to the National Department of Health for implementation support and access to vaccination rates (in particular Tania Van der Merwe), the NICD for support and oversight, and the Network for Genomics Surveillance in South Africa (NGS-SA) for sequence frequencies. Our gratitude to the laboratories, clinicians and data teams in all public and private sector hospitals throughout the country reporting cases and hospitalisation data, who are acknowledged and listed as the DATCOV author group: https://www.nicd.ac.za/diseases-a-z-index/covid-19/surveillance-reports/daily-hospital-surveillance-datcov-report/

## CONTRIBUTORSHIP

WJ, SSAK, CM contributed to literature search. WJ, LB, CC, LO, CM contributed to study design and refining methods of analysis. CM, WJ, SSAK and RW contributed to data analysis, and creation of tables and figures. WJ, SSAK, CC and CM contributed to data interpretation and initial draft. WJ and SSAK drafted the initial manuscript and all other co-authors contributed scientific inputs equally towards the interpretation of the findings and the final draft of the manuscript. WJ, CM, RW and LO have verified the underlying data.

## REFERENCES

1. Abdool Karim SS, Abdool Karim Q. Omicron SARS-CoV-2 variant: a new chapter in the COVID-19 pandemic. Lancet 2021; 38:2126–2128. https://doi.org/10.1016/S0140-6736(21)02758-6

2. Viana R, Moyo S, Anoako DG, Tegally H, Scheepers C, et al. Rapid epidemic expansion of the SARS-CoV-2 Omicron variant in southern Africa. medRxiv, 2021. https://www.medrxiv.org/content/10.1101/2021.12.19.21268028v1 (accessed 22 December 2021)

3. Network for Genomics Surveillance in South Africa (NGS-SA). SARS-CoV-2 Genomic Surveillance Update. https://www.nicd.ac.za/wp-content/uploads/2022/01/Update-of-SA-sequencing-data-from-GISAID-14-Jan-2022_dash_v2-Read-Only.pdf (accessed 17 January 2022).

4. Scott L, Hsiao N, Moyo S, Singh L, Tegally H, et al. Track Omicron’s spread with molecular data. Science, 374 (6574), DOI: 10.1126/science.abn4543 https://www.science.org/doi/epdf/10.1126/science.abn4543?adobe_mc=MCMID%3D30746561878028182403523123834118226244%7CMCORGID%3D242B6472541199F70A4C98A6%2540AdobeOrg%7CTS%3D1639999909

5. Wolter N, Jassat W, Walaza S, Welch R, Moultrie H, et al. Early assessment of the clinical severity of the SARS-CoV-2 omicron variant in South Africa: a data linkage study. The Lancet, 2022. https://doi.org/10.1016/S0140-6736(22)00017-4

6. Updated estimates of the prevalence of SARS-CoV-2 antibodies among blood donors in South Africa. 1 July 2021. https://sanbs.org.za/wp-content/uploads/2016/09/updated-estimates-of-the-prevalence-of-sars-cov-2-antibodies-among-blood-donors-in-south-africa.pdf (accessed 10 December 2021).

7. Madhi S, Kwatra G, Myers JE, Jassat W, Dhar N et al. South African Population Immunity and Severe Covid-19 with Omicron Variant. medRxiv, 2021 https://www.medrxiv.org/content/10.1101/2021.12.20.21268096v1 (accessed 22 December 2021)

8. National Department of Health. Latest Vaccine Statistics. https://sacoronavirus.co.za/latest-vaccine-statistics/ (accessed 14 November 2021).

9. COVID-19 South African Online Portal. https://sacoronavirus.co.za/ (accessed 20 December 2021).

10. Jassat W, Cheryl Cohen, Kufa T, Goldstein S, Masha M, et al. DATCOV: A sentinel surveillance programme for hospitalised individuals with COVID-19 in South Africa, 2020. COVID-19 Special Public Health Surveillance Bulletin. Johannesburg: NICD; 2020. https://www.nicd.ac.za/wp-content/uploads/2020/06/COVID-19-Special-Public-Health-Surveillance-Bulletin-10-June-2020-005.pdf

11. Statistics South Africa. Mid-year population estimates, 2020. Statistical release PO302, Pretoria; 2021. http://www.statssa.gov.za/publications/P0302/P03022020.pdf (accessed 20 December 2021).

12. National Institute for Communicable Diseases. Proposed definition of COVID-19 wave in South Africa. Communicable Diseases Communiqué, November 2021, Vol. 20 (11). https://www.nicd.ac.za/wp-content/uploads/2021/11/Proposed-definition-of-COVID-19-wave-in-South-Africa.pdf (accessed 14 December 2021)

13. Jassat W, Mudara C, Ozougwu L, et al. Difference in mortality among individuals admitted to hospital with COVID-19 during the first and second waves in South Africa: a cohort study. The Lancet Global Health 9 (2021) pp. e1216–e1225. https://doi.org/10.1016/S2214-109X(21)00289-8

14. World Health Organization. Living guidance for clinical management of COVID-19, 23 November 2021. Geneva, 2021. https://apps.who.int/iris/rest/bitstreams/1394399/retrieve

15. Peralta-Santos A, Rodrigues EF, Moreno J, Ricoca V, Casaca P, et al. Omicron (BA.1) SARS-CoV-2 variant is associated with reduced risk of hospitalization and length of stay compared with Delta (B.1.617.2). medRxiv 2021. https://doi.org/10.1101/2022.01.20.22269406 (preprint)

16. Bager P, Wohlfahrt J, Bhatt S, Edslev SM, Sieber RN, et al. Reduced risk of hospitalisation associated with infection with SARS-CoV-2 Omicron relative to Delta: A Danish cohort study. Available at SSRN: https://ssrn.com/abstract=4008930

17. Public Health England; SARS-CoV-2 variants of concern and variants under investigation in England. Technical briefing 34. 14 January 2022. Available: https://assets.publishing.service.gov.uk/government/uploads/system/uploads/attachment_data/file/1046853/technical-briefing-34-14-january-2022.pdf accessed 17 January 2022.

18. Dan JM, Mateus J, Kato Y, Hastie KM, Yu ED, Crotty S, et al. Immunological memory to SARS-CoV-2 assessed for up to 8 months after infection. Science, 371 (6529), eabf4063. DOI: 10.1126/science.abf4063

19. Chan MCW, Hui KPY, Ho J, Cheung M, Ng K, et al. SARS-CoV-2 Omicron variant replication in human respiratory tract ex vivo. https://assets.researchsquare.com/files/rs-1189219/v1/af627c8c-38f1-4a35-9006-4377bfb2decd.pdf?c=1640194833 (preprint)

20. Meng B, Ferreira I, Abdullahi A, Saito A, Kimura I, et al. SARS-CoV-2 Omicron spike mediated immune escape, infectivity and cell-cell fusion. https://www.biorxiv.org/content/10.1101/2021.12.17.473248v2.full (preprint)

21. Bentley EG, Kirby A, Sharma P, Kipar A, Mega DF, et al. SARS-CoV-2 Omicron-B.1.1.529 Variant leads to less severe disease than Pango B and Delta variants strains in a mouse model of severe COVID-19. bioRxiv https://www.biorxiv.org/content/10.1101/2021.12.26.474085v2

22. Diamond M, Halfmann P, Maemura T, Iwatsuki-Horimoto K, ida S, et al. The SARS-CoV-2 B.1.1.529 Omicron virus causes attenuated infection and disease in mice and hamsters. Res Sq 2021;Posted Dec 29: DOI: 10.21203/rs.3.rs-1211792/v1

23. Ryan KA, Watson RJ, Bewley KR, Burton C, Carnell O, et al. Convalescence from prototype SARS-CoV-2 protects Syrian hamsters from disease caused by the Omicron variant. bioRxiv https://www.biorxiv.org/content/10.1101/2021.12.24.474081v1

24. Shinde V, Bhikha S, Hoosain Z, Archary M, Bhorat Q, et al. Efficacy of NVX-CoV2373 Covid-19 Vaccine against the B.1.351 Variant. N Engl J Med 2021;384:1899–909. DOI: 10.1056/NEJMoa2103055

25. Pulliam JRC, van Schalkwyk C, Govender N, von Gottberg A, Cohen C, et al. Increased risk of SARS-CoV-2 reinfection associated with emergence of the Omicron variant in South Africa. medRxiv 2021.11.11.21266068; doi: https://doi.org/10.1101/2021.11.11.21266068 (accessed 18 December 2021)

26. Le Bert N, Tan AT, Kunasegaran K, et al. SARS-CoV-2-specific T cell immunity in cases of COVID-19 and SARS, and uninfected controls. Nature 584, 457–462 (2020). https://doi.org/10.1038/s41586-020-2550-z

27. Abul.Z Raddad LJ, Chemaitelly H, Bertollini R. Severity of SARS-CoV-2 Reinfections as Compared with Primary Infections. N Engl J Med 2021;385;26. DOI: 10.1056/NEJMc2108120

28. Abdool Karim SS, de Oliveira T. New SARS-CoV-2 variants—clinical, public health, and vaccine implications. N Engl J Med 2021;384(19):1866–1868.

29. Rosenberg ES, Dorabawila V, Easton D, Bauer UE, Kumar J, et al. Covid-19 Vaccine Effectiveness in New York State. N Engl J Med 2021. DOI: 10.1056/NEJMoa2116063

30. Collie S, Champion J, Moultrie H, Bekker LG, Gray G. Effectiveness of BNT162b2 Vaccine against Omicron Variant in South Africa. N Engl J Med 2021. https://doi.org/10.1056/NEJMc2119270

31. Cele S, Jackson L, Khan K, Khoury D, Sigal A, et al. SARS-CoV-2 Omicron has extensive but incomplete escape of Pfizer BNT162b2 elicited neutralization and requires ACE2 for infection. medRxiv, 2021. https://www.medrxiv.org/content/10.1101/2021.12.08.21267417v1.full.pdf (accessed 20 December 2021)

